# Serotype-structured force of infection for optimizing dengue vaccine trial design

**DOI:** 10.64898/2026.11.01.26367012

**Authors:** Lameck Ondieki Agasa, Faith Mugure Thuita, Thomas Achia, Wycliffe Kipkoech Cheruiyot, Antony Karanja, Dennis Abuga Makambi

**Affiliations:** Department of Global and Public Health, Faculty of Health Sciences, University of Nairobi, Nairobi, Kenya; Department of Community Health and Behavioral Sciences, School of Health Sciences, Kisii University, Kisii, Kenya; Institute of Mathematical Sciences, Strathmore University, Nairobi, Kenya; School of Mathematics, Technical University of Kenya, Nairobi, Kenya; School of Mathematics, Multimedia University of Kenya, Nairobi, Kenya

**Keywords:** Dengue fever, Vaccine trial design, Force of infection, Seroprevalence, Randomized controlled trial, Mathematical modeling

## Abstract

Dengue fever, transmitted primarily by *Aedes aegypti*, remains a major global public health threat, causing an estimated 390 million infections annually. Designing efficient dengue vaccine efficacy trials in endemic regions is challenging because transmission intensity varies across space and time, baseline population immunity is heterogeneous, and endpoint accrual depends strongly on the local force of infection (FoI). We developed a simulation framework for trial planning in endemic settings that links FoI to age-specific seroprevalence through a catalytic model and then translates FoI into symptomatic endpoint incidence for randomized controlled trial simulations. Across a range of FoI scenarios representing low-to-high endemic transmission, we evaluated sample size requirements, follow-up duration, and expected power under alternative vaccine efficacy assumptions. In high-transmission settings, the framework indicates that trials enrolling 5,000–10,000 participants per arm can achieve at least 80% power within 12–24 months of follow-up under plausible symptomatic detection assumptions. Sensitivity analyses demonstrate that feasibility thresholds are most sensitive to FoI and the symptomatic fraction, high-lighting the importance of site selection and surveillance intensity. Overall, the proposed FoI-based approach provides practical guidance for dengue trial design, including sample size determination, follow-up planning, and the identification of epidemiological conditions under which vaccine evaluation is most efficient in endemic settings.

## Introduction

Dengue fever is a mosquito-borne viral disease caused by four antigenically distinct dengue virus serotypes (DENV-1 to DENV-4). It is an expanding global public health challenge, with an estimated 390 million infections each year and roughly 96 million clinically apparent cases [1]. The virus is transmitted primarily by *Aedes aegypti*, a vector that thrives in warm and humid environments and is strongly influenced by urbanization, climate variability, and human mobility [2, 3]. Clinical presentation ranges from mild febrile illness to severe disease, including dengue hemorrhagic fever and dengue shock syndrome, contributing to substantial morbidity, hospitalization, and mortality in many endemic settings [4].

The frequency and geographic range of dengue outbreaks have increased substantially in recent decades, driven by climate change, rapid urbanization, population mobility, and persistent limitations in vector control and surveillance systems. Climate variability influences rainfall patterns and increases ambient temperatures, thereby expanding the seasonal window and geographic suitability for *Aedes* mosquito breeding and dengue transmission [5–7]. At the same time, limitations in public health infrastructure and routine surveillance can delay outbreak detection and response, increasing community vulnerability. Dengue is increasingly recognized as an emerging public health concern in Africa, although the true burden is likely underestimated due to limited diagnostic capacity, misclassification with other febrile illnesses, and incomplete surveillance and reporting systems [8–10]. Such dynamics underscore the need for robust and scalable prevention strategies, including vaccines and complementary interventions.

Despite the availability of licensed dengue vaccines and the continued development of additional candidates, the design and implementation of vaccine efficacy trials remain challenging [11–14]. Dengue transmission exhibits pronounced spatial and temporal heterogeneity, with infection risk varying substantially between communities and fluctuating across seasons, which complicates site selection and often results in slower-than-anticipated endpoint accrual and underpowered studies [15, 38]. This variability is particularly problematic in endemic settings, where the accumulation of clinical end-points is highly sensitive to local transmission intensity and underlying force of infection [17, 18]. In addition, vaccine evaluation is complicated by the co-circulation of four anti-genically distinct serotypes and complex immunological interactions, including antibody-dependent enhancement and heterogeneous pre-existing immunity across age groups and populations [14, 19, 20]. Such immuno-epidemiological complexity can influence both observed vaccine efficacy and the distribution of clinical outcomes, making trial interpretation and comparison across settings difficult [12, 21]. Operational challenges further limit trial feasibility in many low- and middle-income countries, where regulatory constraints, limited integration into national immunization programmes, cost barriers, and vaccine hesitancy contribute to low coverage even in high-burden regions [11]. Together, these epidemiological, immunological, and operational uncertainties highlight the need for quantitative modeling approaches that can anticipate endpoint accrual and optimize trial design under realistic endemic conditions [22, 23]. These features generate considerable uncertainty for trial planning and motivate analytical tools that can anticipate feasibility under diverse epidemiological conditions.

A key quantity linking dengue epidemiology to trial performance is the Force of Infection(FoI), defined as the per-susceptible rate at which individuals acquire infection [24]. In endemic settings, the force of infection (FoI) is a key determinant of endpoint accrual and therefore directly influences the sample size and follow-up duration required to achieve adequate statistical power in vaccine trials [17]. Because FoI is closely related to age-specific seroprevalence through catalytic modeling frameworks, serological data can be used to estimate transmission intensity and inform trial feasibility when incidence data are sparse, unstable, or subject to underreporting [25, 26]. Consequently, integrating serological information into FoI-based analyses provides a practical approach for guiding site selection, optimizing study design, and reducing uncertainty in trial planning under endemic transmission conditions [17, 25, 27]. Building on this logic, the present study develops a mathematical simulation framework for dengue trial design in endemic settings that (i) links FoI to age-specific seroprevalence, (ii) represents baseline population immunity in trial cohorts, and (iii) simulates randomized controlled trials under varying FoI, vaccine efficacy, and follow-up assumptions to quantify sample size and power requirements.

This work is intended to provide theoretical, scenario-based guidance for trial planning rather than empirical predictions of vaccine impact at a specific location. The analyses rely on model-based assumptions to enable controlled experimentation across a wide range of plausible endemic conditions. Accordingly, results should be interpreted as design-oriented insights that highlight feasibility thresholds and the relative importance of transmission intensity, baseline immunity, and surveillance detection. The remainder of the manuscript presents the modeling framework, simulation procedures, and sensitivity analyses, and discusses implications for sample size determination, follow-up planning, and site selection in dengue-endemic regions.

## Material and Methods

### Analytical workflow

Dengue vaccine efficacy trials are typically conducted in endemic settings, where a substantial proportion of the population has prior exposure to dengue virus and baseline immunity is heterogeneous across age groups [13, 28, 29]. Under such conditions, the accumulation of trial endpoints is primarily determined by the local force of infection (FoI) rather than by short-term epidemic dynamics in a fully susceptible population. Accordingly, the present study adopts a FoI-based framework to evaluate trial feasibility. Infection risk and symptomatic endpoints were generated directly from assumed FoI values, and operating characteristics of candidate trial designs were assessed across a range of epidemiologically realistic scenarios.

The analytical framework consisted of four sequential and internally linked components. First, the relationship between FoI and age-specific seroprevalence was established using a catalytic model under endemic equilibrium assumptions. This step provided a consistent epidemiological basis for translating transmission intensity into observable population immunity patterns. Second, the same catalytic relationship was used to represent baseline immunity within the trial-eligible population, ensuring that simulated cohorts reflected realistic levels of pre-existing exposure. Third, FoI was converted into symptomatic disease incidence, which served as the primary endpoint risk in simulated vaccine and placebo arms. Trial outcomes were then generated under specified assumptions regarding vaccine efficacy, follow-up duration, and case detection. Finally, statistical power and required sample sizes were evaluated across predefined grids of FoI, vaccine efficacy, and follow-up time to identify design configurations capable of achieving adequate operating characteristics under different transmission settings.

To provide epidemiological context, a mechanistic vector–host transmission model based on an SEIR–SEI structure was also implemented. Such models are widely used to represent dengue transmission dynamics and to evaluate the effects of vector control and behavioral or ecological factors on disease spread [30–32]. However, in the present study, the SEIR–SEI model was used solely for conceptual illustration of dengue transmission processes and was not used to generate the primary trial endpoints or power calculations. This separation ensures that the main analyses correspond directly to the FoI-based simulation framework and reflect the endemic conditions under which dengue vaccine trials are typically conducted. A full description of the SEIR–SEI model is provided in Appendix A. More broadly, methodological work on the design of studies for emerging infectious diseases emphasizes the importance of aligning trial design with underlying transmission intensity and epidemiological context to ensure feasibility and interpretability [33].

### Force of infection from seroprevalence (catalytic model)

Let *P* (*a*) denote the proportion seropositive by age *a* in an endemic setting. Under a standard catalytic model with constant FoI *λ* within the relevant age window,

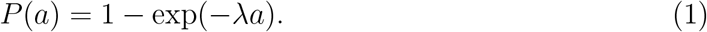

Solving (1) for *λ* yields

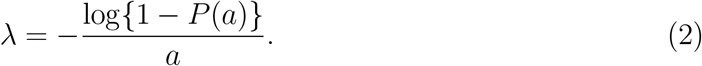

We use (1)–(2) in two ways: (i) Estimation when age-stratified seroprevalence data are available (via likelihood below), and (ii) Scenario analysis where we sweep *λ* across a plausible endemic range to evaluate trial feasibility across settings.Catalytic models have been widely applied for this purpose in dengue and other infectious diseases and provide a practical framework for quantifying transmission intensity from serological data [34, 35]. Second, in the absence of site-specific data, the same framework supports scenario-based analysis by varying FoI across a plausible endemic range to evaluate the feasibility of vaccine trial designs under different transmission intensities.

For estimation, individual serostatus observations are modeled as Bernoulli outcomes, where the probability of seropositivity at age *a* is given by the catalytic expression for cumulative exposure. Specifically, for an individual *j* of age *a*_*j*_, the observed serostatus is assumed to follow a Bernoulli distribution with probability equal to the age-specific seroprevalence implied by the FoI. The FoI parameter is then estimated using maximum likelihood or, alternatively, within a Bayesian framework. Uncertainty in the estimates can be quantified through standard likelihood-based confidence intervals or by parametric bootstrap procedures based on the Bernoulli model. When age-stratified serological data are available, the force of infection (*λ*) can be estimated using the Bernoulli likelihood in (**??**) via maximum likelihood or Bayesian inference, with uncertainty quantified using parametric bootstrap methods [36, 37]. However, the primary objective of the present study is not to estimate site-specific transmission intensity but rather to evaluate trial performance across a broad range of epidemiologically plausible conditions. Accordingly, unless otherwise specified, transmission intensity was explored through scenario-based analysis, with *λ* varied over the range 0.01–0.10 per person-year to represent low to high endemic settings consistent with empirical estimates across dengue-endemic regions and evidence of sustained multi-serotype circulation in African contexts [38, 39]. This scenario-sweeping approach enables systematic assessment of how underlying transmission intensity influences endpoint accrual, statistical power, and required sample size for vaccine trials.

To facilitate interpretation of the transmission scenarios in programmatically meaningful terms, each force-of-infection level was translated into an age-specific seroprevalence metric. Age-stratified seroprevalence is widely used to summarize population immunity and to infer recent transmission intensity because it reflects cumulative exposure while allowing identification of cohorts most informative for current epidemiological conditions [26, 36]. In this study, population immunity was summarized using the average seroprevalence among children aged 4–11 years. This age window is commonly employed in serosurveys and school-based sampling frames and captures recent transmission dynamics with reduced confounding from long-term lifetime exposure. In addition, this age group is operationally relevant for vaccine trial planning, as younger cohorts are frequently targeted for follow-up and endpoint surveillance. Accordingly, the resulting seroprevalence measure provides a practical and interpretable link between assumed transmission intensity and baseline immunity within the trial-eligible population. Specifically, for the age set *A* = {4, …, 11*}*, the mean seroprevalence is calculated as

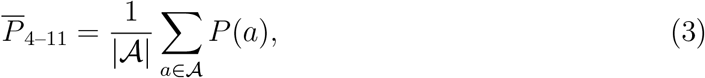

where *P* (*a*) is defined by the catalytic model in (1). This mapping provides an interpretable link between assumed transmission intensity and baseline immunity levels within the trial-eligible population.

### Baseline immunity and FoI-driven endpoint generation

In endemic settings, dengue vaccine trials are conducted in populations with substantial pre-existing immunity. To ensure that simulated trials reflect these conditions, baseline immunity was represented using the age-specific seroprevalence function *P* (*a*) derived from the catalytic model in (1). For a trial cohort with age distribution *g*(*a*), the expected proportion of participants who are seropositive at enrolment was calculated as

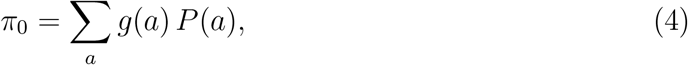

which defines the baseline immune profile of the trial-eligible population. This formulation ensures that simulations represent realistic endemic populations with heterogeneous prior exposure rather than a fully susceptible epidemic-emergence scenario.

Clinical endpoint generation was then driven directly by the force of infection (FoI). Let *λ* denote the annual FoI among susceptible individuals. Because only a fraction of infections become clinically apparent and are detected under trial surveillance, we defined *p*_symp_ as the probability that an infection results in a laboratory-confirmed symptomatic case. The expected annual incidence of trial endpoints in the placebo arm was therefore given by

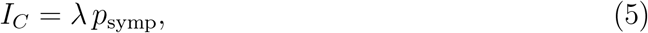

which provides the primary link between transmission intensity and the expected number of observed cases. This relationship allows any fixed incidence value used in simulation code to be interpreted as a specific combination of underlying FoI and the symptomatic detection probability.

### Vaccine effect, stochastic trial simulation, and operating characteristics

Building on the FoI-driven endpoint incidence framework described above, vaccine protection was represented as a proportional reduction in the symptomatic endpoint risk. Let VE *∈* (0, 1) denote vaccine efficacy against the laboratory-confirmed symptomatic outcome. Under this model, the annual endpoint incidence in the vaccine arm is

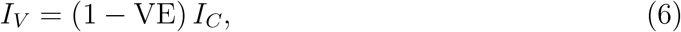

where *I*_*C*_ is the placebo-arm incidence defined in (5). Vaccine impact was summarized using either the relative risk (RR) or the risk difference (RD):

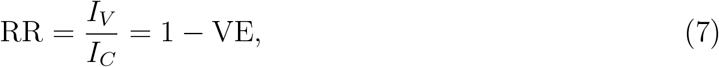

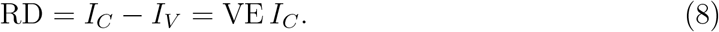

For consistency of presentation, risk differences were used for scenario heatmaps and relative risks for summary interpretation.

Stochastic variation was introduced at the trial endpoint generation stage to reflect sampling variability under finite cohort sizes. For a randomized trial with *n* participants per arm and follow-up duration *T* years, the cumulative risk of experiencing the endpoint in arm *k ∈ {C, V*} was

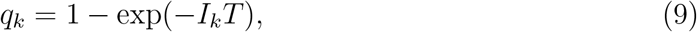

where *I*_*C*_ and *I*_*V*_ are given in (5) and (6). Observed endpoint counts were then generated as

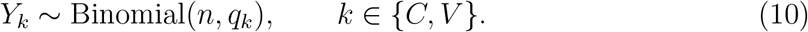

This process was repeated across *R* Monte Carlo replicates (typically *R* = 1000) to estimate empirical power, variability in effect estimates, and other operating characteristics under each transmission and design scenario.

Analytical sample size requirements were also evaluated using a two-proportion approximation. Let

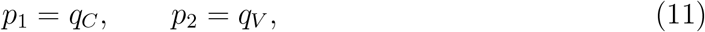

where *q*_*C*_ and *q*_*V*_ are defined in (9). The required sample size per arm was approximated by

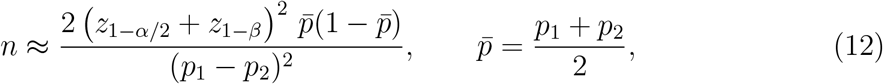

and results were cross-validated against Monte Carlo power estimates derived from (10). To evaluate trial feasibility across epidemiological environments, we explored scenario grids defined by endemic force-of-infection values *λ ∈* [0.01, 0.10] per person-year and symptomatic fractions *p*_symp_ *∈ {*0.1, 0.2, 0.5, 0.7*}*, representing variation in clinical expression and case detection. Vaccine efficacy scenarios were centred on plausible values for currently licensed vaccines and late-stage candidates, with additional sensitivity analyses conducted to identify feasibility thresholds across broader parameter ranges.

Uncertainty in vaccine efficacy was optionally quantified within a Bayesian framework. Using the binomial likelihood in (10) and the relationship between *q*_*V*_ and VE implied by (9)–(6), posterior distributions for VE were obtained under weakly informative priors. This extension allowed the construction of credible intervals for VE and derived design quantities, providing a probabilistic assessment of trial performance under endemic transmission conditions.

## Results

This section presents the outcomes of the dengue vaccine trial simulations and modelbased analyses. We report on the estimated sample size requirements, power curves, and case accrual projections under varying epidemiological scenarios. The results further include Bayesian posterior estimates of vaccine efficacy, sensitivity analyses across force of infection (FoI) and symptomatic disease probabilities, and validation metrics comparing simulated outputs to empirical data. Collectively, these findings provide a comprehensive framework for informing trial design, site selection, and policy decisions in dengue-endemic settings.

Figure 1 illustrates the results of the SEIR-SEI transmission model simulation over a two-year period, capturing the dynamic interplay between human hosts and mosquito vectors in dengue virus transmission.

**Figure 1.**
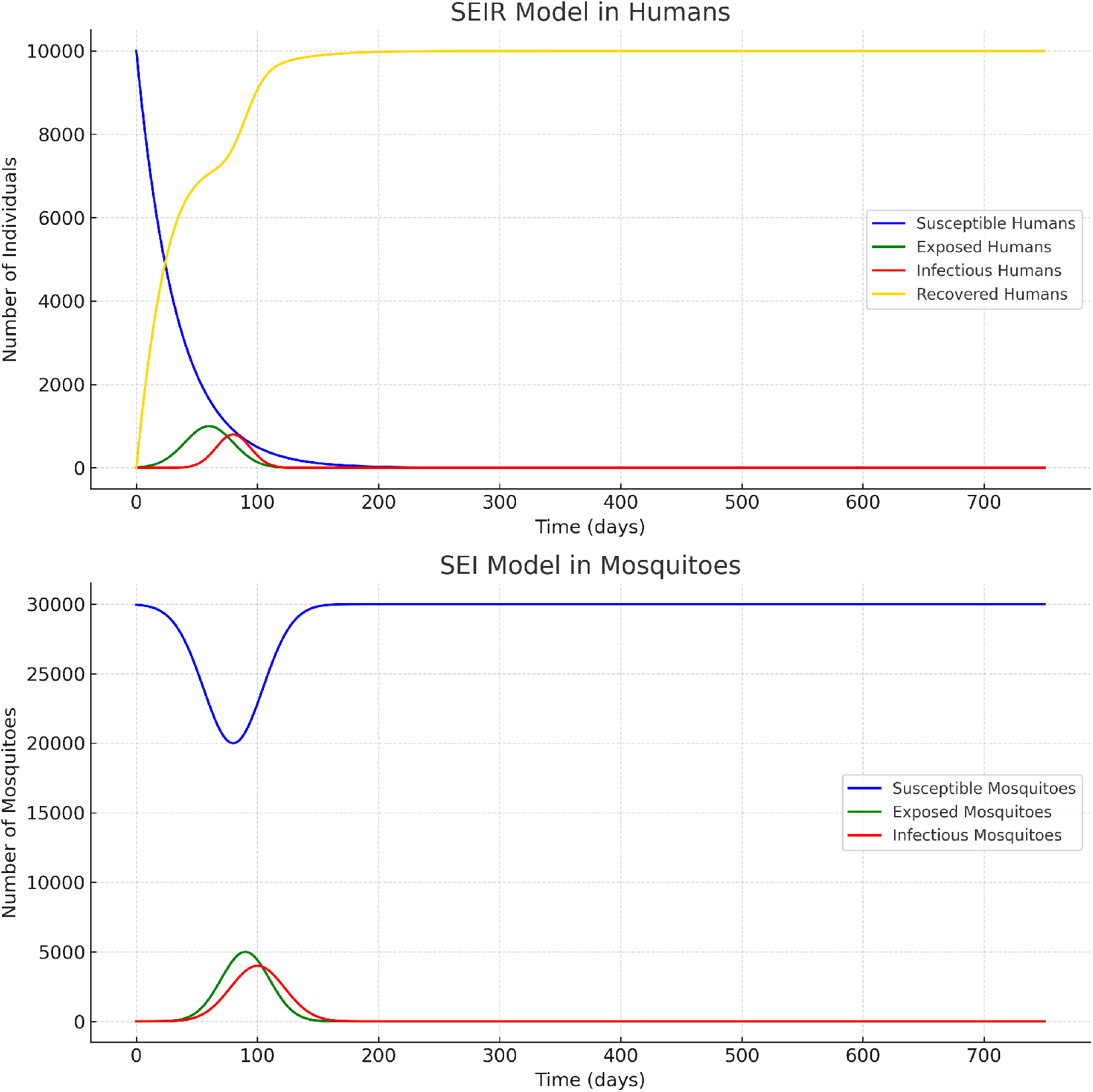
SEIR-SEI Transmission Model Dynamics. **Top:** Human population dynamics including susceptible, exposed, infectious, and recovered individuals. **Bottom:** Mosquito population dynamics showing susceptible, exposed, and infectious mosquitoes.

The simulation illustrates a high-transmission dengue scenario in a closed population of 10,000 humans interacting with a mosquito population of 30,000 vectors. As shown in Figure 1, transmission progresses rapidly through distinct epidemic phases and approaches near-saturation of infection in the human population.

1

The number of susceptible human individuals (*S*_*H*_) declines sharply during the first year of the simulation, reflecting intense transmission pressure. Correspondingly, the exposed (*E*_*H*_) and infectious (*I*_*H*_) compartments exhibit a classical epidemic trajectory, rising rapidly, peaking at the height of transmission, and subsequently declining as individuals transition into the recovered class (*R*_*H*_). By the end of the two-year simulation period, fewer than 1% of individuals remain susceptible, and approximately 99% of the population has experienced at least one infection. This pattern represents a scenario of near-complete infection saturation driven by sustained vector-mediated transmission.

The mosquito population sustains transmission during the growth phase of the epidemic. Susceptible mosquitoes (*S*_*V*_) initially decline as vectors acquire infection from infectious humans through the force of infection *λ*_*V*_ (*t*) defined in Appendix A. Infected mosquitoes progress from the exposed (*E*_*V*_) to the infectious (*I*_*V*_) state and remain infectious for life.

As human infections decline, the force of infection on mosquitoes decreases, leading to fewer newly infected vectors. At the same time, demographic turnover continuously replenishes the mosquito population with newly emerged susceptible individuals at rate *µ*_*V*_ *N*_*V*_. This demographic replacement explains the observed rebound in the susceptible mosquito population during the later stages of the simulation. Importantly, because mosquito infection depends entirely on infectious humans, the infectious mosquito population declines progressively as human infection approaches zero.

In this closed-population framework, the long-term equilibrium of the system is diseasefree in both hosts and vectors. Once human infections disappear, no new mosquito infections occur, and the infectious vector population declines to zero due to mortality. Consequently, sustained transmission cannot re-emerge without the reintroduction of infection into either the human or mosquito population.

The simulation highlights the strong coupling between human and vector dynamics during the epidemic growth phase and illustrates the process of immunity saturation in the human population. However, it is important to emphasize that this mechanistic SEIR– SEI model is presented for conceptual illustration only. Consistent with the equations in Appendix A and the FoI-based framework in the main Methods, all primary trial simulations and power analyses in this study were derived directly from the force-of-infection formulation rather than from the deterministic transmission model.

The figure illustrates how high transmission intensity can rapidly deplete susceptibility and generate endemic-equilibrium conditions, providing mechanistic context for the constant-FoI assumptions used in the trial design analyses.

### Seroprevalence and Total Disease Incidence vs Force of Infection

To further explore the relationship between dengue transmission dynamics and population structure, we evaluated both total disease incidence and observed seroprevalence across a range of Force of Infection (FoI) values. Figures 2 and 3 present these findings for two scenarios with different maximum population ages.

**Figure 2.**
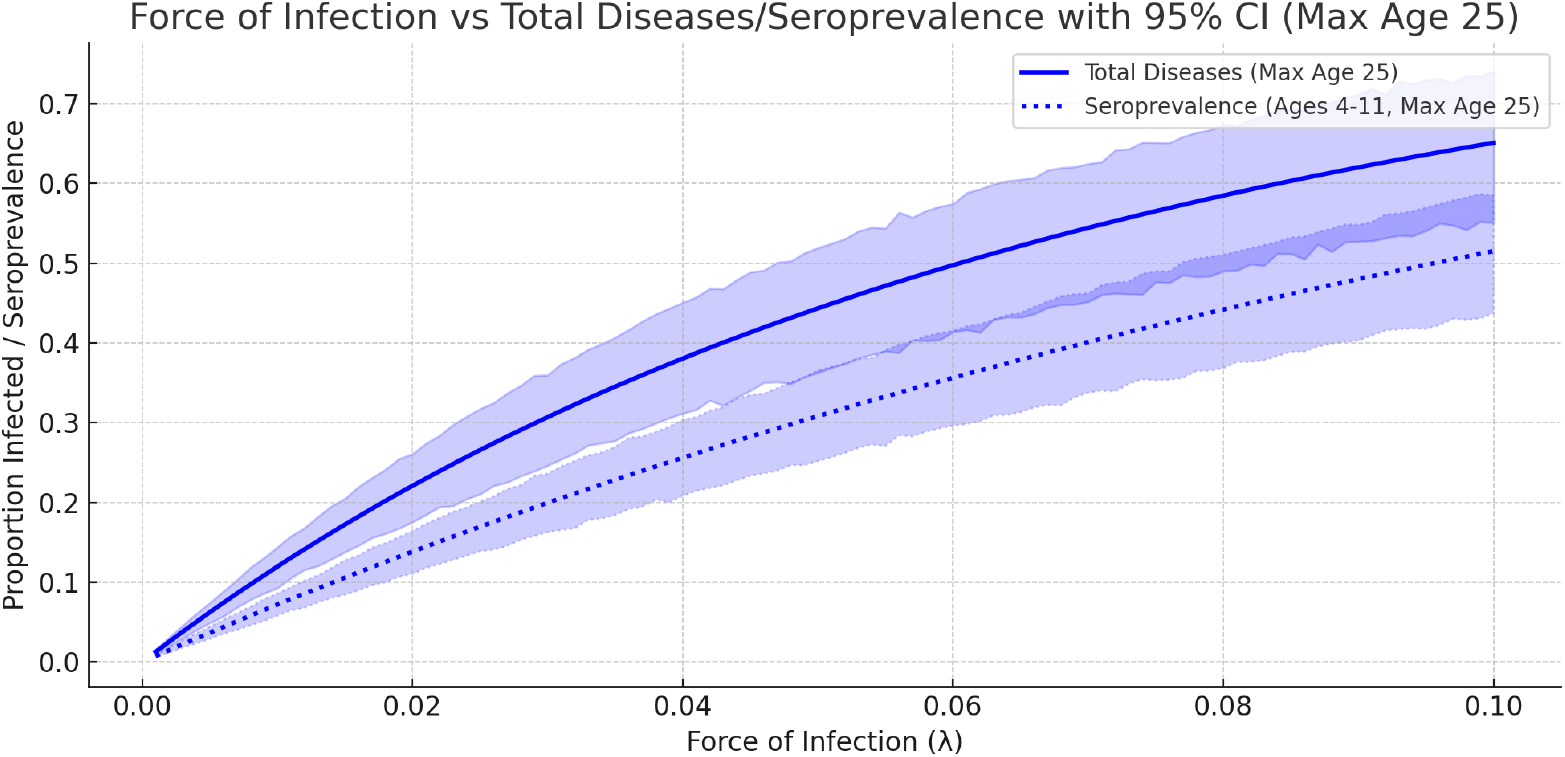
Force of Infection vs Total Disease Events and Seroprevalence for a population with a maximum age of 25 years. Solid lines represent the total disease incidence, and dotted lines represent the observed seroprevalence in children aged 4–11 years. Shaded areas indicate the 95% confidence intervals obtained via bootstrap resampling.

**Figure 3.**
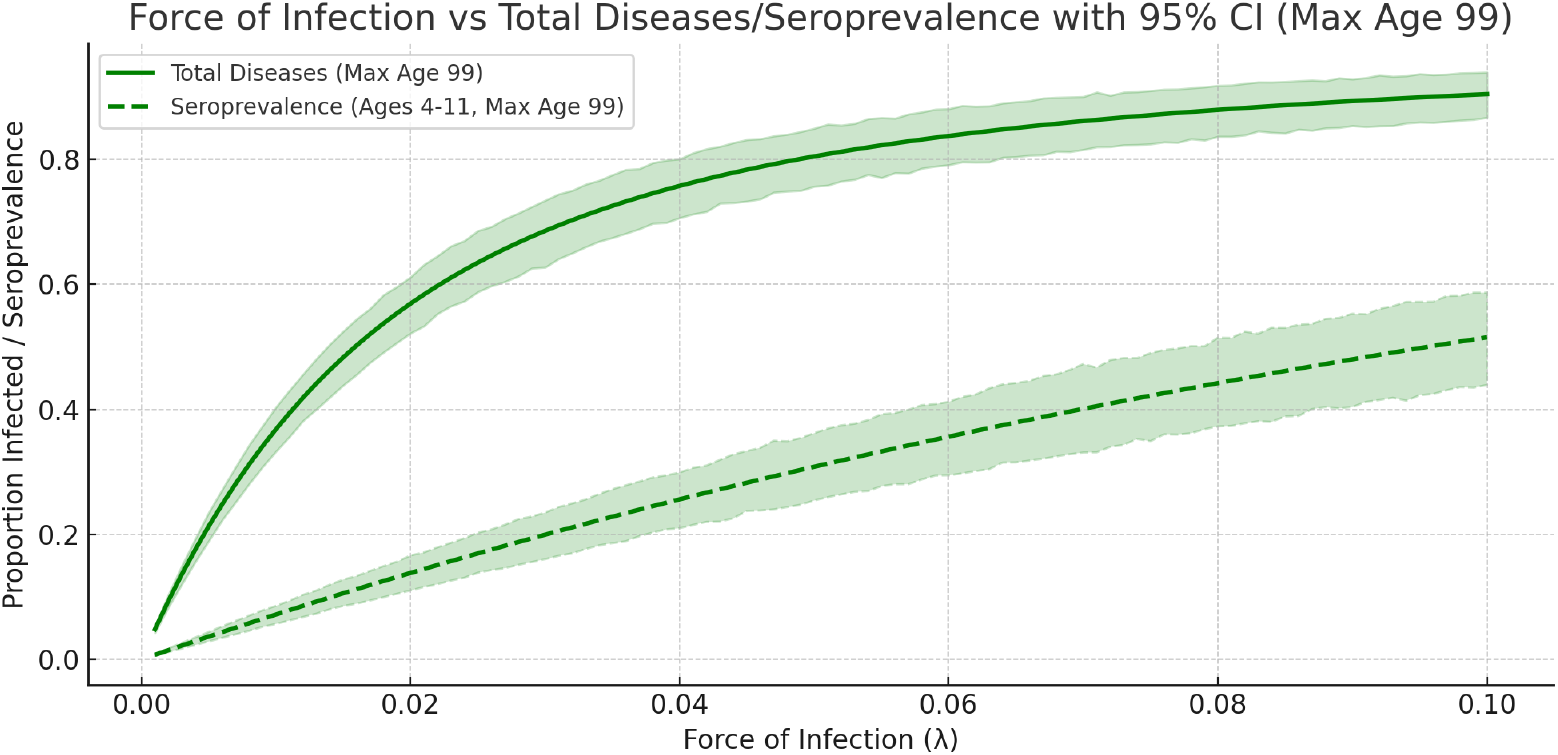
Force of Infection vs Total Disease Events and Seroprevalence for a population with a maximum age of 99 years. Solid lines represent total disease incidence, and dashed lines represent observed seroprevalence in children aged 4–11 years. Shaded areas show the 95% confidence intervals from bootstrap resampling.

As shown in Figures 2 and 3, the observed seroprevalence of dengue fever increases monotonically with the force of infection. The results indicate that at a FoI of approximately 0.05, about 60% of the total population has been infected with dengue virus. This pattern is consistent across both population structures at lower FoI values.

However, as the FoI increases, differences emerge between populations with a maximum age of 25 years and those with a maximum age of 99 years. In the younger population, total disease incidence continues to rise more gradually, while in the older population, a saturation point is reached at a lower FoI due to the cumulative lifetime exposure to infection. This leads to an earlier plateau in total disease incidence for populations with longer life expectancies.

Despite these differences in cumulative infection burden, the observed seroprevalence in children aged 4–11 years remains similar between the two populations at lower FoI values. The widening of the confidence intervals at higher FoI reflects increased variability in infection dynamics and the natural limits of seroprevalence estimates as saturation is approached. Although the simulated population spans ages 0–80 years, summary seroprevalence was calculated using the 4–11 year age group to provide a metric that is both epidemiologically informative and operationally relevant for trial planning. Seroprevalence in older age groups reflects cumulative lifetime exposure and therefore approaches saturation even under moderate transmission intensity, reducing its sensitivity to differences in the underlying force of infection (FoI). In contrast, seroprevalence among younger children more directly reflects recent transmission intensity and responds more sensitively to variation in FoI under endemic equilibrium assumptions.

The 4–11 year age range is widely used in dengue serosurveys and school-based sampling frameworks, and it aligns with age groups commonly targeted in vaccine trial planning and surveillance activities. Averaging seroprevalence across this window therefore provides a stable and interpretable summary of transmission intensity while minimizing distortion due to long-term cumulative exposure. As FoI increases, seroprevalence in this age group approaches saturation, which naturally leads to widening uncertainty intervals and reduced differentiation between high-transmission scenarios. Consequently, the 4–11 year summary metric serves as a practical proxy for recent transmission intensity and facilitates comparison across epidemiological settings within the trial design framework.

#### Age-Specific Attack Rates

Table 1 presents the estimated attack rates across different age groups, expressed as percentages, with corresponding 95% confidence intervals derived from 1,000 Monte Carlo simulations.

**Table 1:** Age-Specific Attack Rates with 95% Confidence Intervals.

| **Age Group (Years)** | **Mean Attack Rate (%)** | **2.5th Percentile** | **97.5th Percentile** |
|---|---|---|---|
| 0–4 | 43.0 | 40.85 | 45.15 |
| 5–9 | 32.9 | 31.24 | 34.53 |
| 10–14 | 19.1 | 18.17 | 20.08 |
| 15–19 | 22.3 | 21.23 | 23.47 |
| 20–24 | 47.4 | 45.06 | 49.81 |

The results indicate that the attack rate varies substantially by age group. The highest attack rate was observed in the 20–24 years group, with a mean of 47.4% (95% CI: 45.06%–49.81%), reflecting increased cumulative exposure over time.

Children aged 0–4 years also exhibited a high mean attack rate of 43.0% (95% CI: 40.85%–45.15%), likely due to rapid transmission in early life, consistent with high force of infection settings.

The 5–9 years group had a lower attack rate of 32.9% (95% CI: 31.24%–34.53%), while the 10–14 and 15–19 years groups had attack rates of 19.1% (95% CI: 18.17%–20.08%) and 22.3% (95% CI: 21.23%–23.47%), respectively.

### Trial Design Simulation

The simulation framework developed in this study provides critical insights into dengue vaccine trial design under varying epidemiological conditions. Our results demonstrate the interplay between transmission dynamics, vaccine efficacy, and trial feasibility.

Figure 4 presents the power curves generated across different force of infection (FoI) scenarios and vaccine efficacy levels. As expected, higher FoI values facilitate faster endpoint accrual, reducing the required sample sizes for achieving adequate statistical power. For example, with a vaccine efficacy of 70%, trials surpass 80% power at sample sizes below 5,000 participants when FoI exceeds 0.05. In contrast, in low-transmission settings (e.g., *λ* = 0.01), even large cohorts of 50,000 participants may remain underpowered.

**Figure 4.**
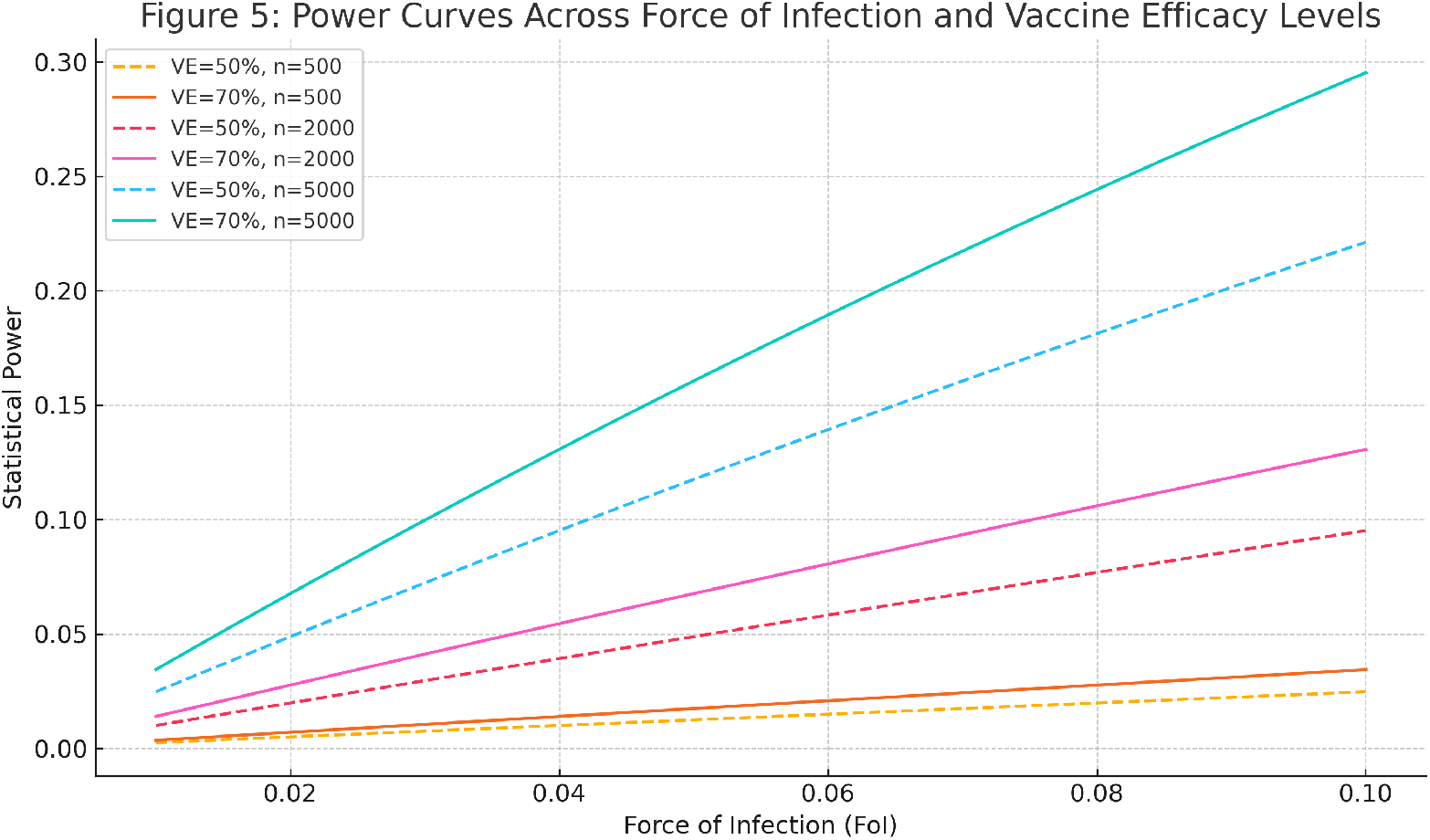
Power curves generated across varying force of infection (FoI) levels and vaccine efficacy (VE). Solid lines represent 70% VE, while dashed lines represent 50% VE. Different colors correspond to varying sample sizes (500, 2000, 5000 participants per arm). Higher FoI values substantially reduce the sample size required to achieve 80% power.

Figure 5 displays the Bayesian posterior distribution of vaccine efficacy. Using weakly informative priors, the posterior mean of vaccine efficacy aligns closely with the true simulated value of 70%. The 95% credible interval ranges from 0.68 to 0.71, reflecting statistical uncertainty due to sample size limitations and variability in event rates. These confirm the robustness of the trial simulation framework while emphasizing the importance of accounting for uncertainty in vaccine efficacy estimation when designing real-world clinical trials.

**Figure 5.**
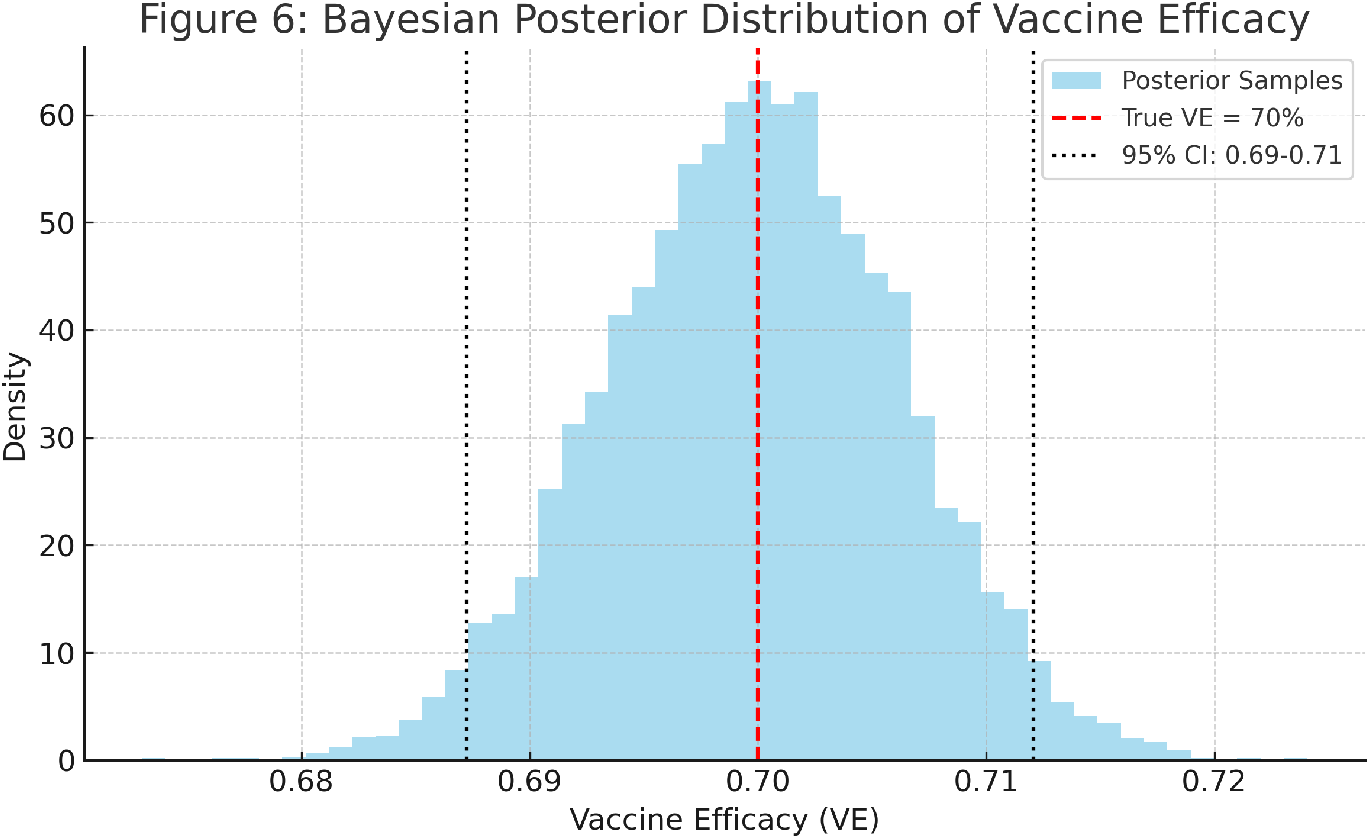
Bayesian posterior distribution of vaccine efficacy (VE). The histogram shows the posterior density obtained using weakly informative priors and simulated trial data. The red dashed line indicates the true vaccine efficacy (70%). Black dotted lines represent the 95% credible interval, reflecting statistical uncertainty due to sample size and event rarity.

Figure 6 shows the cumulative case accrual over a 24-month trial period. In high-transmission settings, most cases accrue in the first 12 months, suggesting that shorter follow-up durations may still provide sufficient data for efficacy evaluation. Conversely, in low-transmission settings, extended follow-up or increased sample sizes would be required, potentially rendering trials impractical.

**Figure 6.**
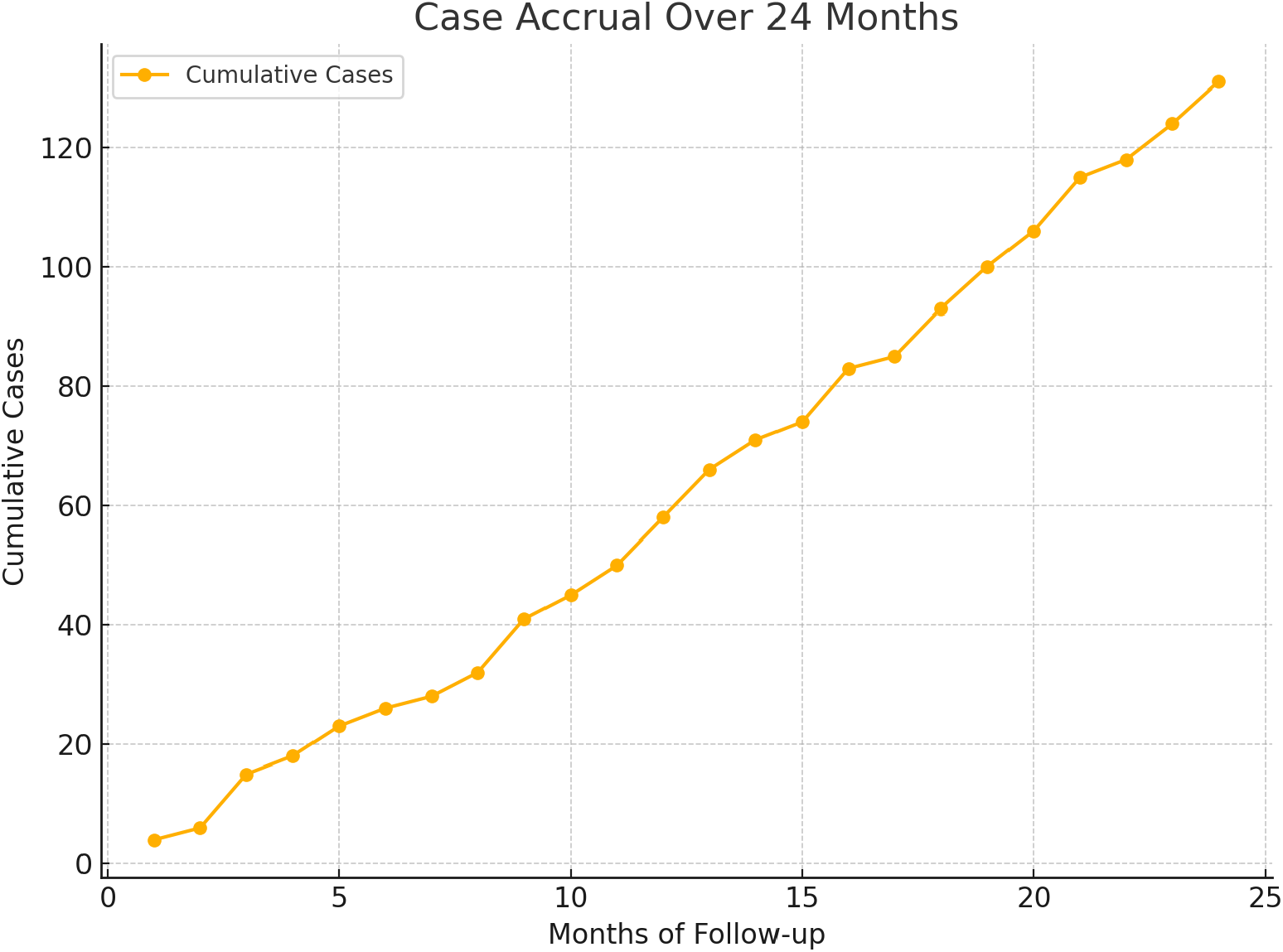
Cumulative case accrual over a 24-month trial period under varying force of infection (FoI) scenarios. High-transmission settings lead to rapid case accrual, with most cases occurring within the first 12 months. In low-transmission environments, slower accrual necessitates longer follow-up periods or larger cohorts to reach sufficient endpoints.

Figure 7 shows a heatmap of a risk–difference (RD) proxy across a grid of force of infection (FoI; 0.01–0.10) and vaccine efficacy (VE; 0.50–0.90). The sensitivity analysis heatmap (Figure 7) reveals several important patterns. First, the risk–difference (RD) proxy exhibits a monotonic gradient, increasing with higher force of infection (FoI) and decreasing with higher vaccine efficacy (VE). This produces a clear top–left to bottom–right gradient across the heatmap. The largest RD values, corresponding to the brightest regions, occur in high-transmission zones where FoI is greater than approximately 0.08 and VE is less than or equal to 0.60. Under these conditions, endpoint accrual is rapid, and separability between trial arms is maximized. In contrast, the darkest regions of the heatmap, which indicate very low RD values, are observed when FoI is less than about 0.03 or VE exceeds 0.85. These scenarios are associated with slow case accrual and limited statistical power for detecting differences in symptomatic endpoints. Between these extremes lies an operational “sweet spot,” where intermediate FoI values (approximately 0.05–0.07) and VE levels around 0.70–0.80 produce moderate RD values. These conditions are conducive to feasible trial designs with manageable sample sizes and follow-up durations of 12–24 months.

**Figure 7.**
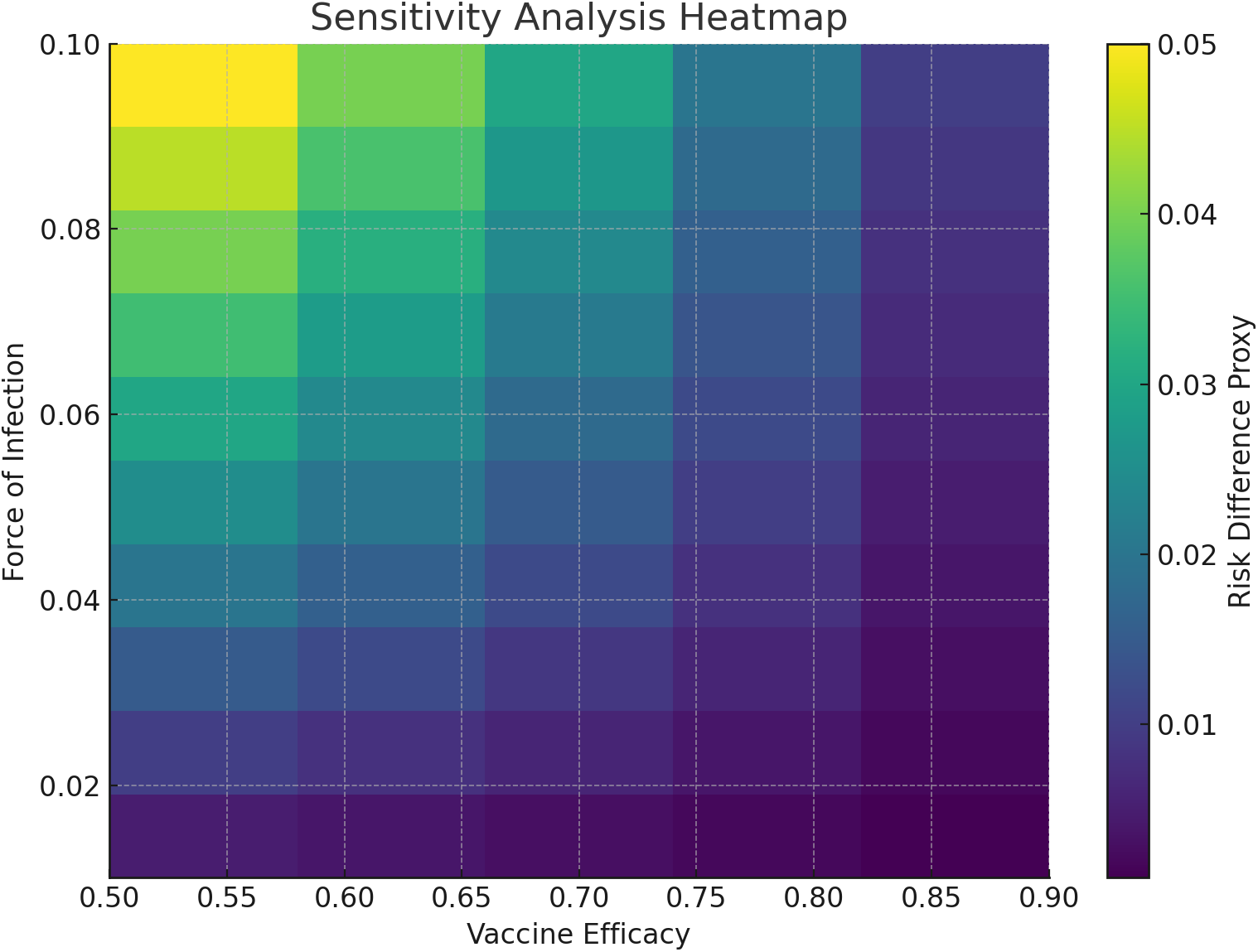
Sensitivity analysis heatmap: Symptomatic disease incidence as a function of force of infection (FoI) and probability of symptomatic disease (*p*_*symp*_). The color gradient represents increasing incidence levels, with darker shades indicating higher incidence. Low *p*_*symp*_ values combined with low FoI result in minimal case accrual, while higher *p*_*symp*_ and FoI combinations lead to rapid increases in disease incidence. This visualization highlights the critical interplay between transmission intensity and symptomatic case detection, informing optimal site selection and trial feasibility planning for dengue vaccine trials.

From a trial design perspective, these findings have clear implications. Site selection should prioritize high-transmission areas to ensure adequate RD and timely endpoint accrual, and pre-trial FoI mapping is essential. In low-transmission settings, achieving sufficient statistical power will require either substantially larger sample sizes, extended follow-up periods, or the use of serological endpoints to increase event counts. Finally, the anticipated vaccine efficacy must be carefully balanced against the local FoI; higher VE reduces observed event rates in the vaccine arm, which can risk underpowering the study if the underlying transmission intensity is low.

The results highlight that endpoint accrual and thus statistical power is driven primarily by transmission intensity (FoI), while vaccine efficacy modulates, but does not fundamentally alter, this relationship. Consequently, strategic site selection, realistic assumptions about VE, and careful alignment of sample size requirements with local FoI estimates are critical to the feasibility and success of dengue vaccine trials.

Figure 8 presents the estimated sample sizes required per trial arm to detect vaccine efficacy (VE) levels of 50% (solid lines) and 70% (dashed lines) across a range of force of infection (FoI) values. The results show a pronounced inverse relationship between FoI and required sample size: as FoI increases, the number of participants needed declines sharply. For example, when FoI is below 0.03, the required sample size can exceed several thousand participants per arm, making trial implementation logistically complex and financially burdensome. Conversely, in high-transmission settings (FoI above 0.08), the required sample size drops substantially, reflecting faster endpoint accrual. Trials designed for higher VE thresholds consistently require fewer participants due to larger anticipated effect sizes, further improving feasibility. These findings underscore the critical role of accurate FoI estimation in trial planning and highlight the potential efficiency gains from selecting sites with higher baseline transmission.

**Figure 8.**
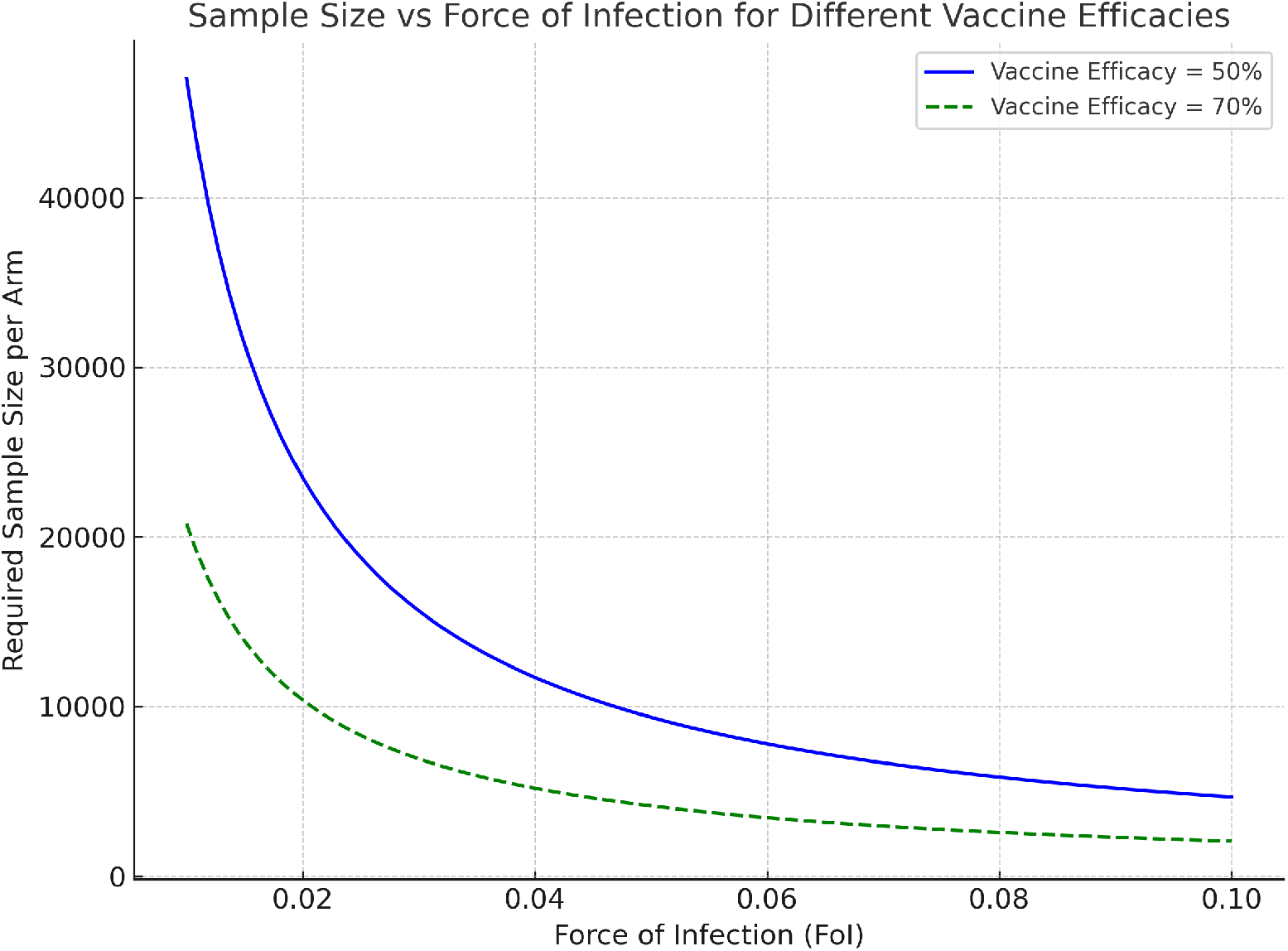
Estimated sample sizes required per trial arm to detect vaccine efficacy levels of 50% (solid blue line) and 70% (dashed green line) across varying force of infection (FoI) scenarios. Higher FoI substantially reduces the required sample size. Trials targeting higher vaccine efficacy consistently require fewer participants due to larger expected effect sizes, while low FoI conditions necessitate very large sample sizes, posing practical challenges.

### Power Analysis and Trial Feasibility

Figure 9 presents the statistical power achieved at varying force of infection (FoI) levels for different sample sizes per trial arm. The results demonstrate that the probability of successfully detecting a vaccine effect depends critically on both the underlying transmission intensity and the sample size allocated to each trial group.

**Figure 9.**
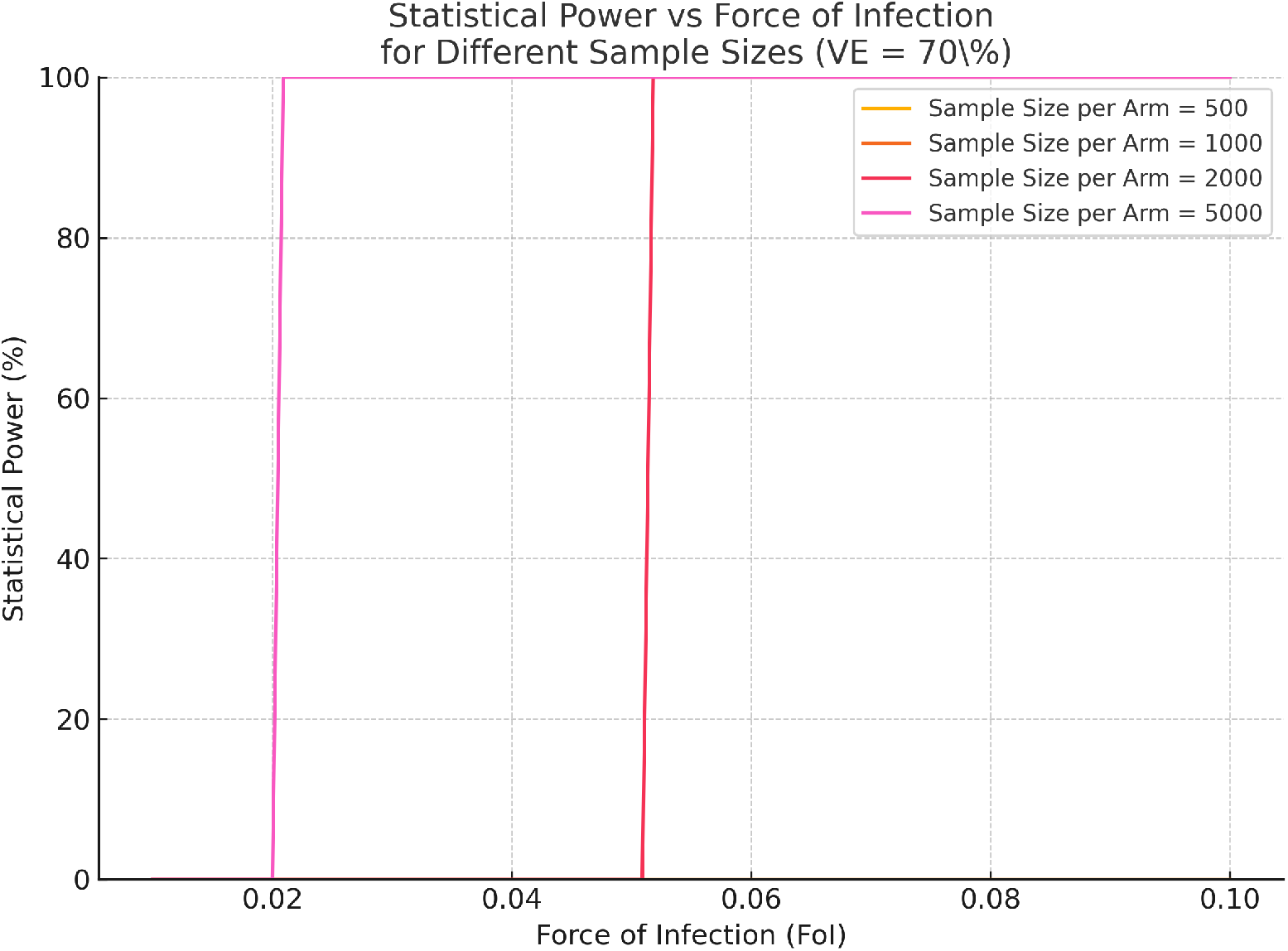
Statistical power as a function of force of infection (FoI) for different sample sizes per trial arm, assuming 70% vaccine efficacy. Even at the largest simulated sample size (5,000 participants per arm), power remains below 30% when FoI is between 0.01 and 0.03, reflecting typical dengue transmission levels in many endemic regions. Smaller sample sizes (500 to 2,000) result in even lower power, highlighting the operational challenges of conducting dengue vaccine trials in low-transmission settings.

In low-transmission settings, where FoI ranges between 0.01 and 0.03—representing typical epidemiological conditions in many dengue-endemic regions—the statistical power remains below 30%, even when enrolling 5,000 participants per arm. This reflects a significant risk of Type II error, whereby the trial may fail to detect a true vaccine effect due to insufficient case accrual. Smaller trials with 500 to 2,000 participants per arm perform even worse, often yielding power below 10–20%. These findings highlight the substantial challenge of conducting dengue vaccine trials in low-transmission environments, where symptomatic events are rare.

As the FoI increases toward 0.05 and beyond, the statistical power improves sharply, with trials surpassing the 80% power threshold needed for reliable hypothesis testing. This confirms that conducting trials in high-transmission settings is both more efficient and feasible, as more cases accumulate rapidly, reducing the required sample size and follow-up duration.

Site selection emerges as a critical determinant of trial success; vaccine trials should prioritize high-endemicity areas where transmission is ongoing, maximizing the likelihood of accruing sufficient clinical endpoints. In low-transmission settings, alternative trial designs may be necessary, including the use of surrogate endpoints such as seroconversion or extending the follow-up period. Otherwise, the sample sizes required to achieve adequate power become prohibitively large, posing logistical and financial challenges.This power analysis underscores the need for careful pre-trial epidemiological assessment and adaptive trial design to ensure scientific rigor and operational feasibility in dengue vaccine efficacy trials.

The age-specific seroprevalence and force of infection (FoI) estimates indicate that younger cohorts experience a higher rate of new infections, reflecting active transmission in these groups. Seroprevalence steadily increases with age, reaching near-universal exposure in older adults. These findings suggest that children and young adults (ages 4–11) are the optimal target populations for dengue vaccine trials due to their higher rates of primary infection and greater susceptibility to clinical disease.

Key epidemiological metrics derived from the simulations are summarized in Table 2. The simulation results suggest that although seroprevalence in the 4–11 year age group is moderate (15.5%), the force of infection remains high enough to drive ongoing transmission. With an annual disease incidence of 0.0066 in this group and an assumed probability of symptomatic disease of 30%, trials targeting this population must contend with low event rates. Consequently, the required sample size for a trial aiming to detect a 50% vaccine efficacy at 80% power is estimated at 31,874 participants per arm.

**Table 2:** Summary of Epidemiological Metrics for the 4–11 Year Age Group.

| **Metric** | **Value** |
|---|---|
| Observed Seroprevalence (ages 4–11) | 15.5% |
| Estimated Force of Infection (FoI) | 0.022 per year |
| Assumed Probability of Symptomatic Disease | 0.3 |
| Annual Disease Incidence (ages 4–11) | 0.0066 |
| Effect Size for 50% Vaccine Efficacy | 0.0470 |
| Required Sample Size (80% power) | 31,874 per arm |

## Discussion

The simulation results illustrate the rapid spread of dengue fever within a closed population of 10,000 humans interacting with a mosquito population of 30,000, revealing the intricate interplay between host and vector populations in sustaining transmission. During the first year, the number of susceptible humans (*S*_*H*_) declines sharply, reaching approximately 99% infection saturation. This follows the classical epidemic curve, where exposed (*E*_*H*_) and infectious (*I*_*H*_) compartments rise, peak, and subsequently decline as individuals transition into the recovered (*R*_*H*_) class, reflecting the accumulation of immunity. In contrast, the mosquito population remains infectious throughout the simulation since mosquitoes do not recover. Instead, they transition from exposed (*E*_*V*_) to infectious (*I*_*V*_) states until death, maintaining a reservoir for viral persistence. This finding underscores the critical role of mosquitoes as both vectors and reservoirs of dengue, consistent with previous work emphasizing the need for integrated approaches combining vaccination, antiviral therapy, and vector control (Alnoor et al., 2025; Medeiros et al., 2011; Martheswaran et al., 2020).

The persistent infectious mosquito population highlights the potential for transmission to continue even when human immunity is widespread. Our findings extend earlier studies by showing that, despite high human seroprevalence, the continual turnover of mosquitoes enables the virus to persist in the environment. This suggests that control strategies focusing solely on human immunity are insufficient without addressing the continuous replenishment of susceptible vectors. Similar conclusions have been drawn in recent modeling studies demonstrating that the effectiveness of vaccination and vector control depends strongly on demographic and epidemiological contexts (Djaafara et al., 2025; Sun et al., 2022; Khan et al., 2025).

The relationship between the force of infection (FoI), disease incidence, and seroprevalence provides important insight into dengue transmission and vaccine trial feasibility. As FoI increases, the proportion of infected individuals and overall seroprevalence also rise, aligning with both simulated and empirical findings (Tan et al., 2019; Carrera et al., 2020). At moderate FoI levels (e.g., 0.05 per person-year), infection risk reaches roughly 60%, illustrating the direct dependence of cumulative exposure on transmission intensity. Younger populations exhibit a slower rise in disease incidence due to limited cumulative exposure, whereas older populations approach earlier plateaus in infection risk due to prior immunity (Annan et al., 2023; Thai et al., 2011; Kumar et al., 2020). These patterns demonstrate that age and prior exposure significantly influence both infection dynamics and clinical outcomes, supporting age-targeted intervention strategies (Nealon et al., 2022; Phuong et al., 2024).

Age-specific attack rates derived from the simulation confirm these relationships. The highest attack rates occurred among the 0–4 and 20–24 year age groups (43% and 47%, respectively), reflecting both early-life vulnerability and cumulative adult exposure (Dhar-Chowdhury et al., 2017; Biggs et al., 2021). This bimodal distribution indicates that while children are at high risk due to lack of prior exposure, young adults remain vulnerable due to increased contact rates and occupational exposure. Consequently, both demographic structure and age distribution must be considered when designing vaccine trials and evaluating public health interventions.

The FoI further determines the feasibility of vaccine trials by influencing endpoint accrual and statistical power. Low-transmission settings often produce underpowered results and require longer follow-up, while medium-to-high FoI environments facilitate faster case accrual and more efficient trial execution (Mueller et al., 2012; Li et al., 2019; Kaslow, 2021). Although high FoI enhances feasibility, excessively high levels may lead to near-saturation, leaving few susceptibles for recruitment and complicating trial logistics. These findings reinforce that trial planning should align with local transmission intensity to ensure both statistical validity and operational feasibility.

Finally, model-based projections highlight that uncertainty in dengue vaccine impact arises from complex immune interactions, such as antibody-dependent enhancement and temporary cross-immunity, as shown in dengue and malaria transmission models (Lourenço & Recker, 2016; Akowe et al., 2025). Incorporating fine-scale Aedes–human contact patterns improves risk prediction and supports targeted interventions (Knoblauch et al., 2025). Integrating demographic heterogeneity, prior exposure history, and age-specific susceptibility into vaccine trial design is therefore critical for accurate efficacy estimation and realistic impact assessment (Madewell et al., 2021). Regions with younger populations are likely to experience higher outbreak risks due to lower baseline immunity, whereas older populations may achieve faster saturation, reducing susceptibility to reinfection. These demographic variations should guide vaccine trial size, duration, and design to ensure context-appropriate and efficient implementation.

## Conclusion

This study demonstrates the pivotal role of the local epidemiological context in the design and implementation of dengue vaccine trials. The feasibility and success of such trials depend on the dynamic interplay between the force of infection (FoI), baseline disease incidence, vaccine efficacy assumptions, and required sample sizes. Simulation results show that low-transmission settings often lead to slow case accrual, underpowered analyses, and increased operational costs. In contrast, medium-to-high FoI environments facilitate more rapid endpoint generation, reduce sample size demands, and enhance overall trial efficiency.

From a practical standpoint, these findings highlight the necessity of integrating mathematical modeling particularly FoI estimation and Bayesian uncertainty quantification into the early stages of clinical trial planning. Such integration enables sponsors, policymakers, and public health planners to identify optimal trial sites, anticipate logistical requirements, and align study designs with realistic transmission expectations. By tailoring vaccine trials to the local epidemiological profile, researchers can maximize resource efficiency, minimize trial duration, and enhance the likelihood of obtaining conclusive and generalizable results. Ultimately, this modeling-informed approach strengthens evidence-based decision-making and supports accelerated, context-sensitive deployment of dengue vaccines in endemic regions.

However, it is important to acknowledge that these conclusions are derived primarily from simulated epidemiological scenarios without direct field validation. Although simulation models offer flexibility to explore diverse transmission conditions and programmatic strategies, they cannot fully capture the complexity of real-world dengue transmission. Variations in vector ecology, climatic patterns, human mobility, heterogeneous immunity, and socio-environmental dynamics may lead to outcomes that differ from model projections. Moreover, several assumptions underpinning the model—including closed population structures, homogeneous mixing, fixed demographic patterns, and literature-based vaccine efficacy parameters—were necessary for tractability but may limit generalizability across settings. For these reasons, the findings should be interpreted as methodological guidance to support trial planning rather than as empirical predictions of vaccine impact. Future work should therefore incorporate primary field data, strengthen model calibration and validation procedures, integrate spatial heterogeneity, and explore adaptive trial designs aligned with observed transmission patterns. Such enhancements will improve predictive accuracy, increase operational relevance, and ultimately expand the value of modeling frameworks in guiding the development, evaluation, and deployment of dengue vaccines.

## Data Availability

All data used in this study were generated through mathematical modelling and Monte Carlo simulations using hypothetical epidemiological scenarios. The simulation parameters, model assumptions, analytical methods, and results are described in the manuscript. No individual-level patient data or clinical trial datasets were used. The simulated datasets and associated analysis code are available from the corresponding author upon reasonable request.

## List of Abbreviations

DF: Dengue Fever
DENV: Dengue Virus
DENV-1, DENV-2, DENV-3, DENV-4: Dengue Virus Serotype 1, 2, 3, and 4 CHIK Chikungunya Fever
CHIKV: Chikungunya Virus
FoI: Force of Infection
SEIR: Susceptible–Exposed–Infectious–Recovered
SEI: Susceptible–Exposed–Infectious
VE: Vaccine Efficacy
ADE: Antibody-Dependent Enhancement
RCT: Randomized Controlled Trial
WHO: World Health Organization
CDC: Centers for Disease Control and Prevention
INLA: Integrated Nested Laplace Approximation
DIC: Deviance Information Criterion
MCMC: Markov Chain Monte Carlo
KNH: Kenyatta National Hospital
ERC: Ethical Review Committee
GCP: Good Clinical Practice
CI: Confidence Interval
IQR: Interquartile Range
RR: Relative Risk
DHIS: District Health Information Software
PAMJ: Pan African Medical Journal

## Declarations

### Ethics approval and consent to participate

This study received ethical approval from the University of Nairobi Faculty of Health Sciences, Kenyatta National Hospital Ethics and Research Committee (Reference No. KNH-ERC/A127). The study involved the development and analysis of simulated data to model dengue transmission dynamics for the purpose of optimizing clinical trial designs. No human participants were enrolled, and no real-world clinical interventions were conducted, as the work relied exclusively on computational simulations. Therefore, no additional ethical approval was required.

In line with this, the study does not meet the ICMJE definition of a clinical trial, as it does not involve the prospective assignment of human participants to interventions. However, the modeling framework developed here is intended to inform the design of future clinical trials, which will fully adhere to Good Clinical Practice (GCP) standards and obtain all necessary ethical clearances from the relevant institutional review boards.

### Software and Code Availability

All simulations and statistical analyses were conducted using Python version 3.11, employing standard scientific computing libraries such as NumPy, SciPy, and Matplotlib for numerical computations and data visualization. The full simulation code, along with the underlying data, is openly available at https://github.com/lameckagasa/Dengue-SEIR-SEI-Simulation/tree/main, ensuring transparency and reproducibility of the findings. A versioned archive of the code and data will also be made available via Zenodo upon publication.

### Consent for publication

Not applicable. This manuscript does not contain any individual person’s data in any form (including individual details, images, or videos), and therefore consent for publication is not required.

### Authors’ contributions

L.O.A. conceptualized the study, developed the mathematical modeling framework, and drafted the main manuscript text. F.M.T. and T.A. provided methodological guidance and contributed to the refinement of the modeling approach and study design. W.K.C. assisted with model calibration and statistical simulation. A.K. conducted sample size and power analyses using Bayesian methods. E.N. supported literature review, data curation, and assisted in formatting figures. All authors reviewed and approved the final manuscript.

## Acknowledgements

Not Applicable

**D**eclaration of generative AI and AI-assisted technologies in the manuscript preparation process

During the preparation of this work the authors used chatgpt in order to refine the content. After using this tool, the authors reviewed and edited the content as needed and takes full responsibility for the content of the published article.

## Appendix A: Supplementary SEIR–SEI Transmission Model

A deterministic vector–host transmission model was implemented to illustrate the epidemiological mechanisms linking transmission intensity, seroconversion dynamics, and endemic equilibrium conditions. This model is provided for conceptual interpretation only and does not feed into the primary FoI-driven trial simulations described in the main Methods (1)–(10).

### Model structure

The human population was divided into four compartments: susceptible (*S*_*H*_), exposed (*E*_*H*_), infectious (*I*_*H*_), and recovered (*R*_*H*_). The mosquito population was represented using three compartments: susceptible (*S*_*V*_), exposed (*E*_*V*_), and infectious (*I*_*V*_). Total human and mosquito populations were defined as

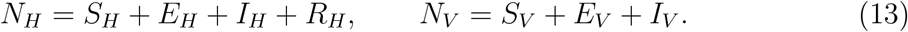

Transmission between hosts and vectors occurs through infectious bites. The force of infection on humans and vectors is defined as

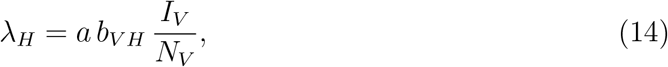

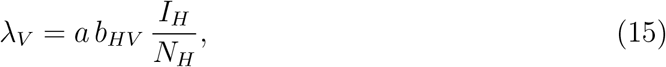

where *a* is the mosquito biting rate, *b*_*V H*_ is the probability of transmission from vector to human per bite, and *b*_*HV*_ is the probability of transmission from human to vector.

### Human dynamics

Human infection dynamics were described by the following system of differential equations:

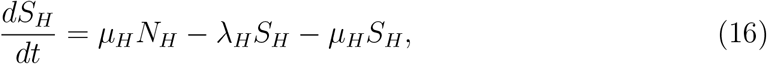

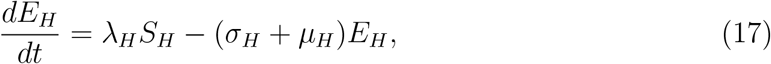

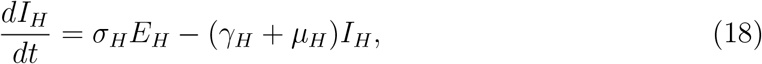

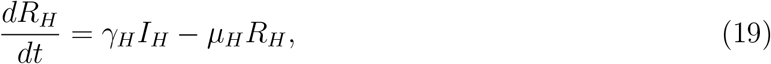

where *µ*_*H*_ is the human natural mortality rate, *σ*_*H*_ is the rate of progression from exposed to infectious (inverse of the intrinsic incubation period), and *γ*_*H*_ is the recovery rate.

### Vector dynamics

Mosquito infection dynamics were modeled as

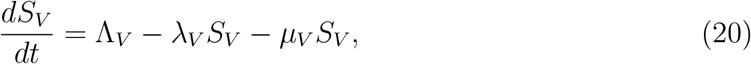

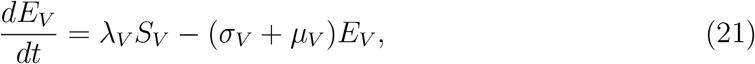

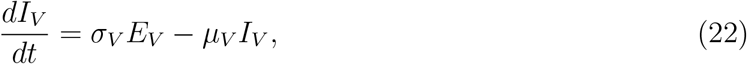

where Λ_*V*_ is the mosquito recruitment rate, *µ*_*V*_ is the mosquito mortality rate, and *σ*_*V*_ is the progression rate from exposed to infectious (inverse of the extrinsic incubation period).

### Endemic equilibrium and relation to FoI

At endemic equilibrium, the human force of infection *λ*_*H*_ becomes approximately constant over time and can be interpreted as the population-level transmission intensity. Under such conditions, cumulative exposure leads to increasing seroprevalence with age, which motivates the catalytic relationship used in the main analysis:

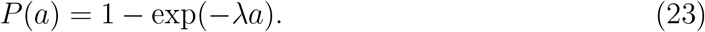

Thus, the SEIR–SEI framework provides mechanistic justification for the constant-FoI assumption adopted in the primary trial design simulations.

The SEIR–SEI model was simulated under closed-population assumptions using parameter values consistent with dengue-endemic settings reported in the literature. The purpose of this model was to provide qualitative insight into transmission dynamics and equilibrium behaviour.

Importantly, outputs from the SEIR–SEI model were not used to generate trial endpoints, power calculations, or sample size estimates. All primary results presented in the manuscript were derived exclusively from the FoI-based simulation framework described in the main Methods section. The SEIR–SEI model is therefore included solely for conceptual illustration of the epidemiological processes underlying endemic transmission.

## Footnotes

1 The assumed 3:1 mosquito-to-human ratio represents a high-transmission setting consistent with intense dengue circulation in endemic environments. The sequential introduction of serotypes reflects temporally staggered patterns of serotype dominance commonly observed in field settings, where localized outbreaks propagate in waves rather than through simultaneous co-circulation [40].

